# Association of puberty timing with Type 2 diabetes: Systematic review and meta-analysis

**DOI:** 10.1101/19000521

**Authors:** Tuck Seng Cheng, Felix R. Day, Rajalakshmi Lakshman, Ken K. Ong

## Abstract

**OBJECTIVE:** We aimed to systematically review published evidence on the association between puberty timing and Type 2 diabetes or impaired glucose tolerance (T2D/IGT), *with* and *without* adjustment for adiposity, and to estimate its potential contribution to the burden of T2D.

**RESEARCH DESIGN AND METHODS:** We searched PubMed, Medline and Embase databases for publications until February 2019 on the timing of any secondary sexual characteristic in boys or girls in relation to T2D/IGT. Inverse-weighted random-effects meta-analysis was used to pool reported estimates and meta-regression to explore sources of heterogeneity.

**RESULTS:** Twenty eight observational studies were identified. All assessed age at menarche (AAM) in women (combined N=1,228,306); only one study additionally included men. In models *without* adjustment for adult adiposity, T2D/IGT risk was higher per year earlier AAM (relative risk (RR)=0.91, 95% confidence interval (CI)=0.89-0.93, 11 estimates, n=833,529, *I*^2^=85.4%) and for early versus later menarche (RR=1.41, 95% CI=1.28-1.55, 23 estimates, n=1,185,444, *I*^2^=87.8%). Associations were weaker but still evident in models adjusted for adiposity (AAM: RR=0.97 per year, 95% CI=0.95-0.98, 12 estimates, n=852,268, *I*^2^=51.8%; early menarche: RR=1.19, 95% CI=1.11-1.28, 21 estimates, n=890,583, *I*^2^=68.1%). Associations were stronger among Caucasians than Asians, and in populations with earlier average AAM. The estimated population attributable risk of T2D in UK Caucasians due to early menarche, unadjusted and adjusted for adiposity, was 12.6% (95% CI=11.0-14.3) and 5.1% (95% CI=3.6-6.7), respectively.

**CONCLUSIONS:** A substantial proportion of T2D in women is attributable to early menarche timing. This will increase in light of global secular trends towards earlier puberty timing.

## INTRODUCTION

Puberty is the transitional period from childhood to adulthood when physiological and physical changes relating to sexual maturation occur to attain fertility. The onset of puberty (Tanner stage 2) is indicated by the appearance of breast buds in girls, genital development in boys, and pubic hair growth in both sexes (1,2). In the latter period of puberty (at Tanner stage 3 or 4), girls experience first menstruation, namely menarche (3) and boys experience voice break (4). Within populations, timing of puberty varies widely by sex and between individuals. The normal age at onset of puberty ranges from 8 to 13 years in girls and from 9 to 14 years in boys (5,6) and age at menarche (AAM) continues to decline worldwide (5,7-9).

Puberty timing has been widely examined in relation to health outcomes, including Type 2 diabetes (T2D) which is increasingly prevalent worldwide (10). An earlier systematic review and meta-analysis showed that early menarche was associated with higher T2D risk (11). That review identified 10 relevant publications (315,428 participants) dated until the end of 2013 and included only two studies in non-Western settings (both were from China) (11), which did not allow for comparisons between regions. There have been several very large Asian studies published subsequently (12,13). More importantly, that previous meta-analysis analyzed only effect estimates adjusted for body mass index (BMI) (11). As BMI was invariably measured in adults, rather than in childhood, it may be considered as a mediator between puberty timing and T2D, rather than simply a confounder, although BMI, overweight and obesity track from early childhood to adulthood (14,15). Comparisons of the associations between AAM and T2D both *with* and *without* adjustment for adiposity would be informative. Furthermore, a recent study from China reported that the association between AAM and incident diabetes differed by year of birth, with a stronger association observed in women who were born in more recent decades (12). Such potential effect modifications were not investigated in the previous meta-analysis (11).

Here, we describe a systematic review and meta-analysis to evaluate the association between puberty timing and T2D and/or impaired glucose tolerance (T2D/IGT), with and without adjustment for adiposity, in both women and men. We also assessed study-design-related factors that could explain the heterogeneity between study estimates. Finally, we describe, to our knowledge, the first estimate of the potential contribution of early menarche timing to the population burden of T2D.

## RESEARCH DESIGN AND METHODS

### Data sources and searches

We searched online databases (i.e., PubMed, Medline and Embase) until 28 February 2019. The search terms were: i) terms or measures related to puberty timing (e.g. puberty, menarche, voice break, Tanner); and ii) terms or measures related to diabetes (e.g. diabetes, glucose, insulin, glycated haemoglobin); and iii) terms related to epidemiological studies (based on guidelines from Scottish Intercollegiate Guidelines Network) (16). Further details of the search strategy are shown in Supplemental Table 1. All identified papers were screened by title and abstract, and if considered potentially relevant, the full texts were read for inclusion decision. Any uncertainty about the eligibility of a particular paper was resolved through discussion between authors (T.S.C. and K.K.O.). We also reviewed papers included in the previous systematic review (11) and reference lists of our included papers to identify relevant papers. The present study was registered in the International prospective register of systematic reviews (PROSPERO Registration Number: CRD42019124353) and the protocol is available at: http://www.crd.york.ac.uk/PROSPERO/display_record.php?ID=CRD42019124353.

### Study Selection

Published papers were included in the present systematic review if they reported: i) any measure of puberty timing either reported in childhood or adulthood (pubertal onset: age at breast or genital development, or Tanner stage 2 pubic hair) (1,2); pubertal completion: AAM, voice breaking, and ii) T2D/IGT assessed by self-reported physician diagnosis, fasting plasma glucose, oral glucose tolerance test and/or glycated haemoglobin. Other inclusion criteria were: any epidemiological study in women or men and published in full reports in English.

### Exclusion criteria

We excluded studies that analysed populations with specific diseases such as breast cancer, polycystic ovary syndrome, Turner syndrome, premature adrenarche and Type 1 or 2 diabetes, as well as animal studies.

### Data Extraction

Data from eligible studies for systematic review were extracted by one author (T.S.C.); a 20% sample was independently extracted by a second author (R.L.), blinded to the original dataset, which was verified (100% agreement) by a third author (K.K.O).

Extracted information included first author, publication year, sample size, study population and ethnicity, year at enrolment, ages at puberty and outcome assessments, mean AAM, number of cases, definition of outcome, types of outcomes (prevalent or incident T2D/IGT cases), risk estimates with corresponding confidence intervals (CI), definitions of early puberty and its reference category, and variables controlled for in multivariable models. Specifically, for meta-analysis, we selected i) risk estimates for T2D/IGT per year later AAM as a continuous variable (i.e., dose-response relationship) and ii) risk estimates for T2D/IGT in the earlier AAM category compared to the middle or older AAM category (i.e., categorical relationship). We distinguished between estimates from models adjusted for potential confounders (but non adiposity) and estimates from models adjusted for adiposity indicators (usually BMI or waist circumference, or preferentially both). If a study reported estimates for multiple outcomes, we prioritised risk estimates for combined T2D/IGT, followed by T2D only and IGT only, and included estimates for only one such outcome per study.

For those studies that reported risk estimates for T2D/IGT per year earlier (rather than later) AAM (17), we calculated the reciprocals to produce risk estimates per year later AAM. Similarly, for those studies that reported risk estimates for T2D/IGT in an older (rather than earlier) AAM category (12,18-21) compared to an earlier AAM category as the reference, we calculated the reciprocals to produce risk estimates in the earlier AAM category compared to the older AAM category as the reference. For simplicity, we considered odds ratios and hazard ratios to be similar estimates of the relative risk (RR).

### Data synthesis and analysis

To summarize the association between AAM and T2D/IGT, inverse-variance weighted random-effects models were performed. Estimates from models *with* and *without* adjustment for adiposity indicators were considered separately. Heterogeneity between studies was quantified by the inconsistency index (*I*^2^) (*I*^2^<50%, 50–75%, and >75% indicated mild, moderate, and high heterogeneity, respectively). Potential sources of heterogeneity were evaluated using meta-regression analyses. Publication bias was evaluated using visual inspection of funnel plots and Egger’s regression asymmetry test. Sensitivity analyses by the trim-and-fill and leave-one-out methods were performed. Statistical analyses were performed using the “metafor” package in R software. P values <0.05 were considered to indicate statistically significance.

Based on the causal assumption that AAM affects T2D/IGT risk, the population attributable risk for T2D/IGT due to early menarche among British women was calculated using the formula: 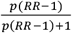, where *p* is the prevalence of early menarche (defined as <12 years) in the large population-based UK Biobank study (22) and RR is the pooled risk estimate among Caucasians.

### Quality assessment

The Newcastle-Ottawa Quality Assessment Scale for cohort studies (23) was used to assess the quality of each study included in the systematic review. Criteria for each item in the assessment scale were defined according to the present research topic before study quality assessments were performed. For longitudinal studies of incident T2D/IGT and longitudinal studies which assessed puberty timing in adolescence and early adulthood and subsequent prevalent T2D/IGT, all 8 items were applied (maximum score of 9). For cross-sectional studies of prevalent T2D/IGT, only 6 items (maximum score of 7) were used (presence of T2D/IGT at baseline, and follow-up duration were not relevant).

## RESULTS

### Study characteristics

Study selection is summarised in Figure 1. The search strategy identified 6155 records. After removing duplicates and non-relevant studies based on titles and abstracts, 49 texts were selected for full-text reading and finally 28 studies were deemed eligible for inclusion in the review. All 10 studies included in the previous review (11) and studies in the reference lists of included studies were found by our search strategy.

**Figure 1.**
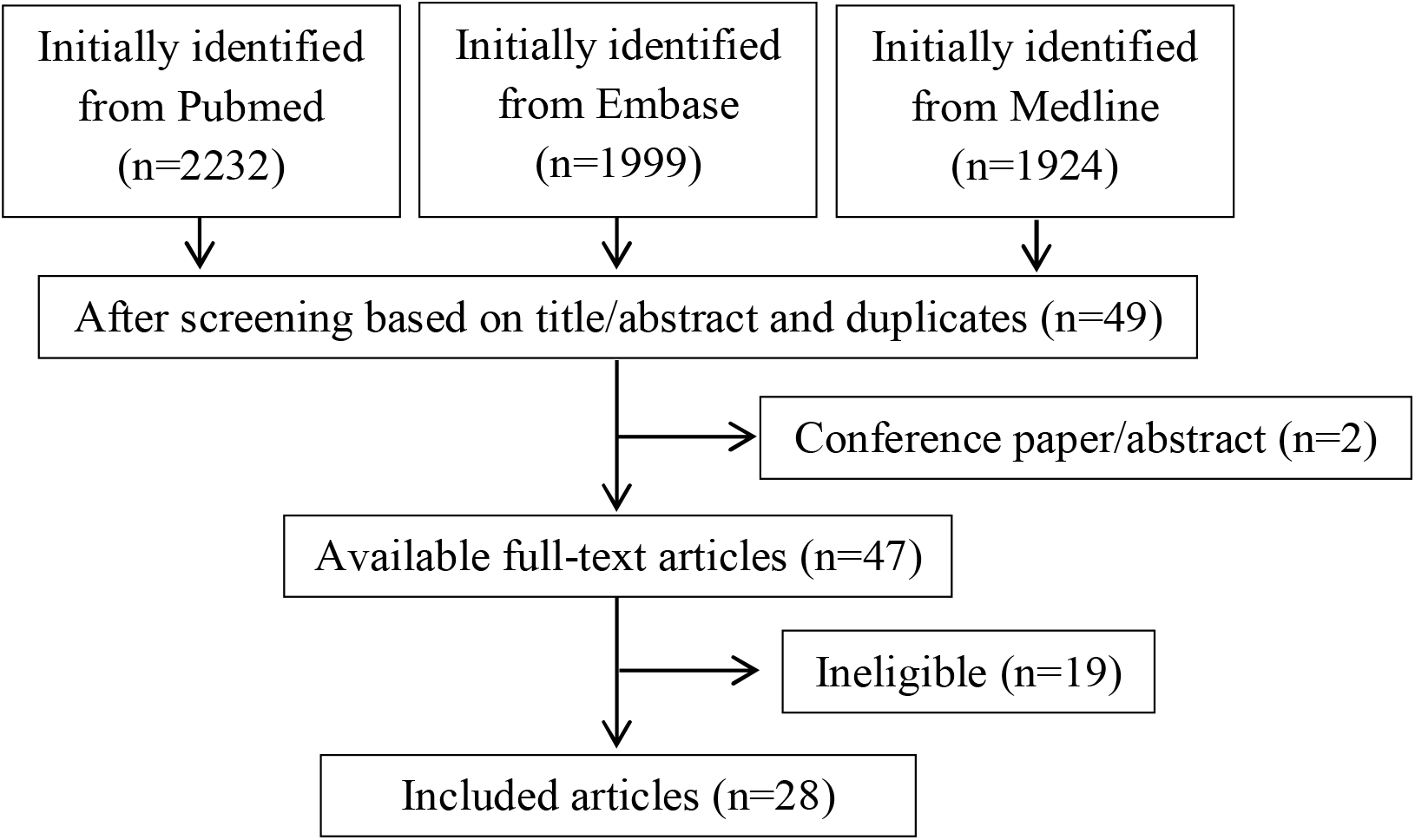
Flowchart of study selection

Tables 1 and 2 (and Supplemental Tables 2 and 3) show the characteristics of the included studies by prevalent and incident cases of T2D/IGT, respectively. Of the 28 included studies, all assessed AAM in women (combined N=1,228,306) and only one additionally analysed age at voice breaking in men (22). The assessment of puberty timing was conducted during mid-late adulthood in most studies (mean ages ranging 35-70 years), except during adolescence in one study (24) and in early adulthood (age <25 years) in two studies (17,25). All were observational studies and one additionally included a Mendelian randomization analysis (13). Nine studies were conducted among Caucasians (18,19,22,24-29), 13 studies among Asians (12,13,20,21,30-38) and 6 studies among multi-ethnic populations (Caucasian, Hispanic, Asian, African-American and Latino) (17,39-43). Fourteen studies examined prevalent T2D (13,18,19,22,24,25,28,32,34-37,41,43), 2 prevalent IGT (21,30), 3 prevalent T2D/IGT (26,31,33), 8 incident T2D (12,17,20,27,29,38,39,42), and one prevalent and incident T2D (40). The definitions of T2D/IGT varied across studies and 4 studies excluded participants with potential Type 1 diabetes based on age at diagnosis (22,24,40,41). The adiposity indicators adjusted for in 25 studies were mostly BMI alone (n=19) (17-21,24-29,33,35,37-39,41-43), followed by both BMI and waist circumference (n=4) (12,32,34,40), waist circumference alone (n=1) (30) and body composition (n=1) (22). Early menarche was defined as AAM <12 years in 9 studies (17,22,27-29,36,39-41) and <14 years in 13 studies (12,18,20,21,30-35,37,38,42), while the reference category of AAM was defined as AAM ≥12 years in 12 studies (22,27-29,31,33-35,39-42) and ≥14 years in 10 studies (12,17,18,20,21,30,32,36-38). Furthermore, the reference category of AAM was the middle category in 12 studies (22,27-29,32-34,37,39-42) and the oldest category in 10 studies (12,17,18,20,21,30,31,35,36,38).

**Table 1.**
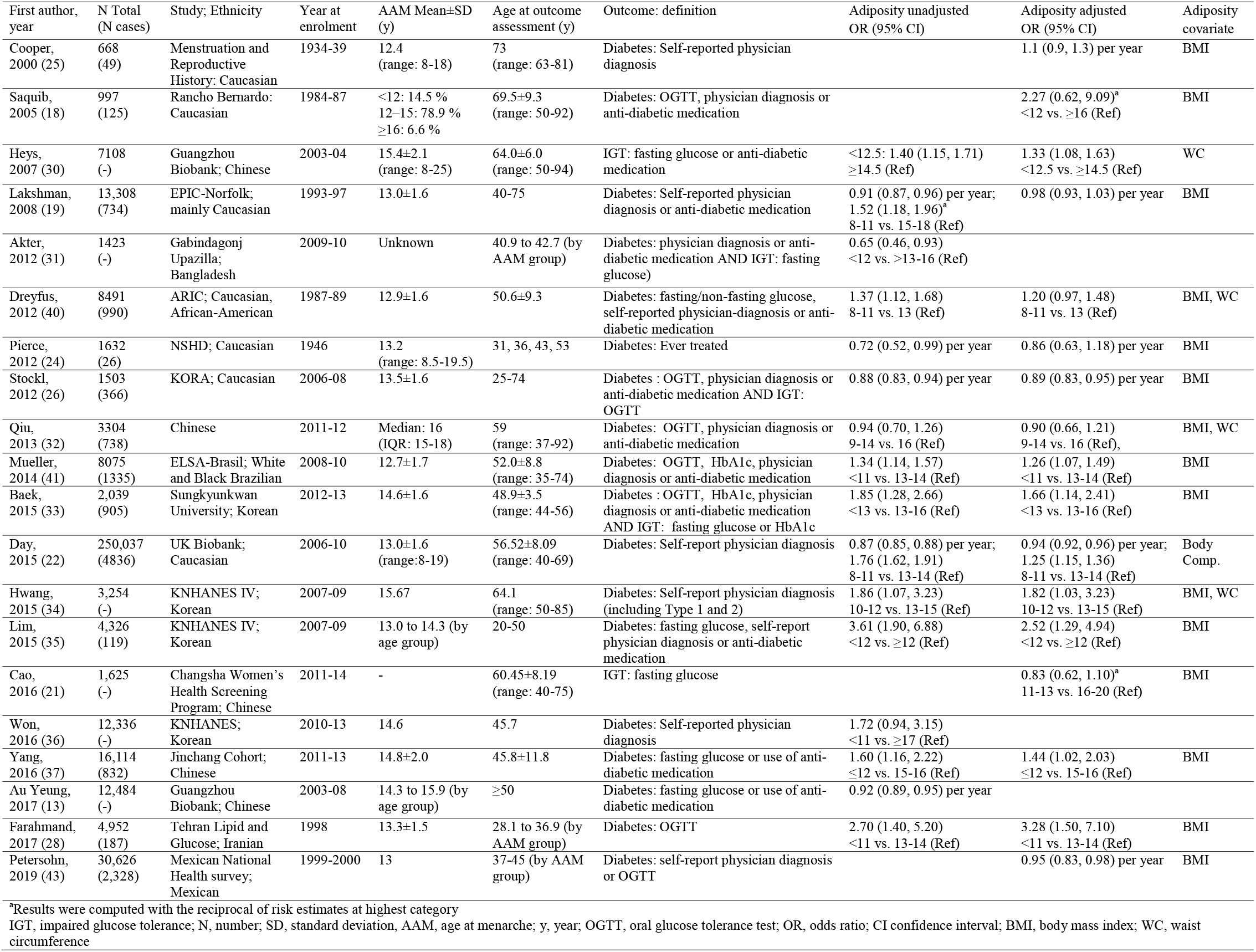
Summary of eligible studies of prevalent diabetes/IGT.

**Table 2.** Summary of eligible studies of incident diabetes/IGT.

| First author, year | Total N (N cases) | Study; Ethnicity | Year at enrolment | AAM (years) | Age at outcome assessment (y) | Outcome: definition | Adiposity unadjusted OR (95% CI) | Adiposity adjusted OR (95% CI) | Adiposity covariate |
| --- | --- | --- | --- | --- | --- | --- | --- | --- | --- |
| He, 2010 (39) | 101,415 (7,963) | Nurses' Health; Multi-ethnic | 1980 | - | 63.5 | Diabetes: OGTT; ≥1 diabetes symptom; anti-diabetic medication | 0.94 (0.92, 0.95) per year; 1.21 (1.13, 1.31) ≤11 vs.13 (Ref) | 0.99 (0.97, 1.01) per year; 1.02 (0.95, 1.10) ≤11 vs. 13 (Ref) | BMI |
|  | 100,547 (2,739) | Nurses' Health II; Multi-ethnic | 1991 | - | 47.4 |  | 0.88 (0.86, 0.91) per year; 1.50 (1.34, 1.69) ≤11 vs.13 (Ref) | 0.97 (0.94, 1.00) per year; 1.15 (1.02, 1.29) ≤11 vs. 13 (Ref) | BMI |
| Conway, 2012 (20) | 69,385 (1,831) | Shanghai Women's Health; Chinese | 1997-2000 | - | 60.1±2.0 | Diabetes: OGTT or anti-diabetic medication | 0.95 (0.92, 0.98) per year; 1.35 (1.14, 1.59)<sup>a</sup> 8-13 vs. 17-26 (Ref) | 0.98 (0.95, 1.01) per year; 1.14 (0.95, 1.33)<sup>a</sup> 8-13 vs. 17-26 (Ref) | BMI |
| Dreyfus, 2012 (40) | 7,501 (755) | ARIC; Caucasian, African-American | 1987-89 | 12.9±1.6 | 56.8±8.0 | Diabetes: fasting/non-fasting glucose, self-reported physician-diagnosis or anti-diabetic medication | 1.27 (1.02, 1.58) 8-11 vs. 13 (Ref) | 1.18 (0.95, 1.47) 8-11 vs. 13 (Ref) |  |
| Elks, 2013 (27) | 10,903 (4,242) | EPIC-InterAct; Caucasian | 1991 | 13.14±1.58 | 52 | Diabetes: Health record confirmed self-reported physician diagnosis | 0.89 (0.86, 0.93) per year; 1.70 (1.48, 1.94) 8-11 vs. 13 (Ref) | 0.96 (0.91, 1.01) per year; 1.42 (1.18, 1.71) 8-11 vs. 13 (Ref) | BMI |
| Dreyfus, 2015 (17) | 1,970 (271) | CARDIA; Caucasian, African-American | 1985 | 12.6±1.5 (range: 8-16) | 50 (range: 42-59) | Diabetes: OGTT or anti-diabetic medication | 0.93 (0.86, 1.00) per year<sup>b</sup>, 1.61 (1.09, 2.37) 8-11 vs. 14-17 (Ref) | 0.90 (0.86, 0.94) per year<sup>b</sup>, 1.33 (0.90, 1.96) 8-11 vs. 14-17 (Ref) | BMI |
| LeBlanc, 2017 (42) | 124,379 (11,262) | Women's Health Initiative; Multi-ethnic | 1993-98 | - | (Follow-up: 12.2±4.2) | Diabetes: self-report diagnosis, use of anti-diabetes medication | 1.14 (1.08, 1.20) <12 vs. 12 (Ref) | 1.01 (0.95, 1.06) <12 vs. 12 (Ref) | BMI |
| Yang, 2018 (12) | 270,345 (5,391) | China Kadoorie Biobank; Chinese | 2004-08 | 15.4±1.9 | (Follow-up: 7) | Diabetes: Health record | 0.96 (0.94, 0.97) per year; 1.33 (1.24, 1.44)<sup>a</sup> 13 vs. ≥18 (Ref) | 0.98 (0.97, 1.00) per year | BMI, WC |
| Pandeya, 2018 (29) | 126,721 (4,073) | InterLACE; mainly Caucasian | 1985-09 | 13.1 (range:8-20) | 56.1±11.4 | Diabetes: self-reported physician diagnosis or Health records | 1.63 (1.40, 1.89) ≤10 vs. 13 (Ref) | 1.18 (1.02, 1.37) ≤10 vs. 13 (Ref) | BMI |
| Nanri, 2019 (38) | 37,511 (513) | Japan Public Health Center-based Study; Japanese | 1990, 1993 | 14.7±1.9 | (Follow-up: 10) | Diabetes: Health record confirmed self-reported physician diagnosis | 1.09 (0.83, 1.43)<sup>a</sup> ≤13 vs. ≥16 (Ref) | 1.01 (0.76, 1.33)<sup>a</sup> ≤13 vs. ≥16 (Ref) | BMI |

<sup>a</sup>Results were computed with the reciprocal of risk estimates at highest category
<sup>b</sup>Results were computed with the reciprocal of risk estimates per year early age at menarche
IGT, impaired glucose tolerance; N, number; SD, standard deviation, AAM, age at menarche; y, year; OGTT, oral glucose tolerance test; OR, odds ratio; CI confidence interval; BMI, body mass index; WC, waist circumference

From models *without* adjustment for adiposity, most studies (n=20/24) reported a statistically significant association with higher T2D/IGT risk for earlier menarche (12,13,17,19,20,22,24,26-30,33-35,37,39-42) or earlier voice breaking (22); only 3 reported no association (32,36,38) and one study reported that earlier menarche was associated with *lower* T2D/IGT risk (31). From models *with* adjustment for adiposity, some studies (n=11/24) reported a statistically significant association with higher T2D/IGT risk for earlier menarche (12,22,26,28-30,33-35,37,41) or younger voice breaking (22), but not other studies (n=11) (17-21,24,25,32,38,40,42) and two studies reported inconsistent findings between dose-response and categorical AAM models (27) or between sub-cohorts (39).

### Quality assessment

More than half of studies of prevalent T2D/IGT (n=11 studies) scored 6/7, followed by 5/7 (n=4), 7/7 (n=3) and 5/9 (n=2) (Supplemental Table 4). Longitudinal studies of incident T2D/IGT were rated 9/9 (n=5) or 8/9 (n=4) (Supplemental Table 5).

### Meta-analysis results

All 28 studies on AAM and T2D/IGT in women were included in the meta-analysis. Similar findings were observed between pooled estimates for T2D only and IGT only (Supplemental Figure 1 and 2). To maximise power, we therefore prioritised risk estimates for combined T2D/IGT (3 studies), followed by T2D only (23 studies) and IGT only (2 studies).

Figure 2 shows the continuous association between AAM and T2D/IGT. From models without adjustment for adult adiposity, pooled analysis of 11 estimates from 10 studies showed that earlier AAM was associated with higher T2D/IGT risk (RR=0.91 per year, 95% CI=0.89-0.93; n=833,529; Figure 2a). This association was weaker but still evident in models with adjustment for adiposity (pooled analysis of 12 estimates from 11 studies: RR=0.97 per year, 95% CI=0.95-0.98; n=852,268; Figure 2b). Similar findings were obtained in subgroup analyses by prevalent or incident T2D/IGT. Heterogeneity between studies was high in estimates without adjustment for adiposity (*I*^2^=85.4%) and moderate in estimates with adjustment for adiposity (*I*^2^=51.8%).

**Figure 2.**
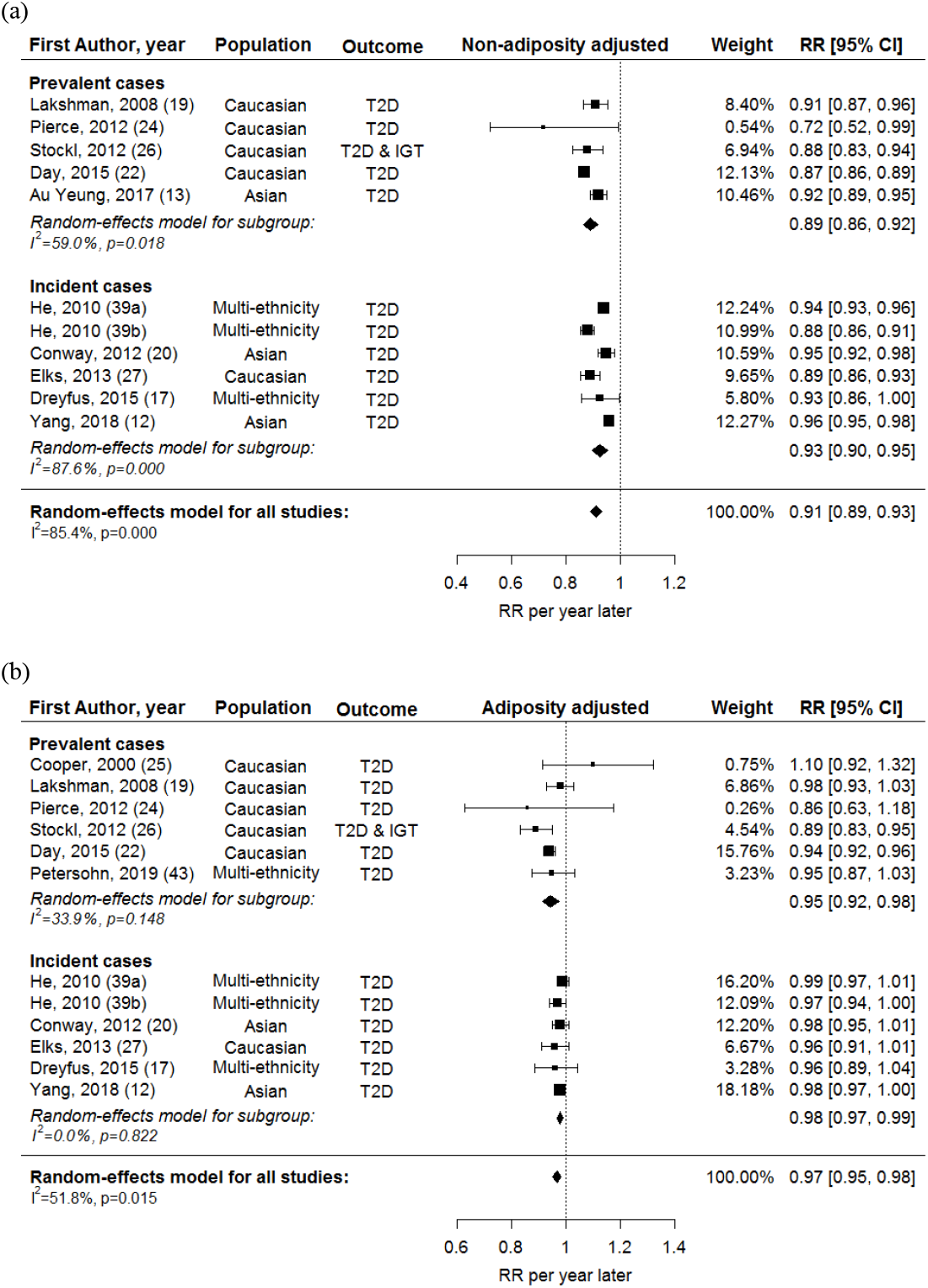
Forest plots of the association between AAM (continuous variable) and T2D/IGT, (a) without and (b) with adjustment for adiposity.

Figure 3 shows the categorical association between early versus later menarche with T2D/IGT. From models without adjustment for adult adiposity, pooled analysis of 23 estimates from 21 studies showed that early menarche was associated with higher T2D/IGT risk (RR=1.41, 95% CI=1.28-1.55; n=1,185,444; Figure 3a). This association was weaker but still evident in models with adjustment for adiposity (pooled analysis of 21 estimates from 19 studies: RR=1.19, 95% CI=1.11-1.28; n=890,583; Figure 3b). Similar findings were obtained in subgroup analyses by prevalent or incident T2D/IGT. Heterogeneity between studies was high in estimates without adjustment for adiposity (*I*^2^=87.8%) and moderate in estimates with adjustment for adiposity (*I*^2^=68.1%).

**Figure 3.**
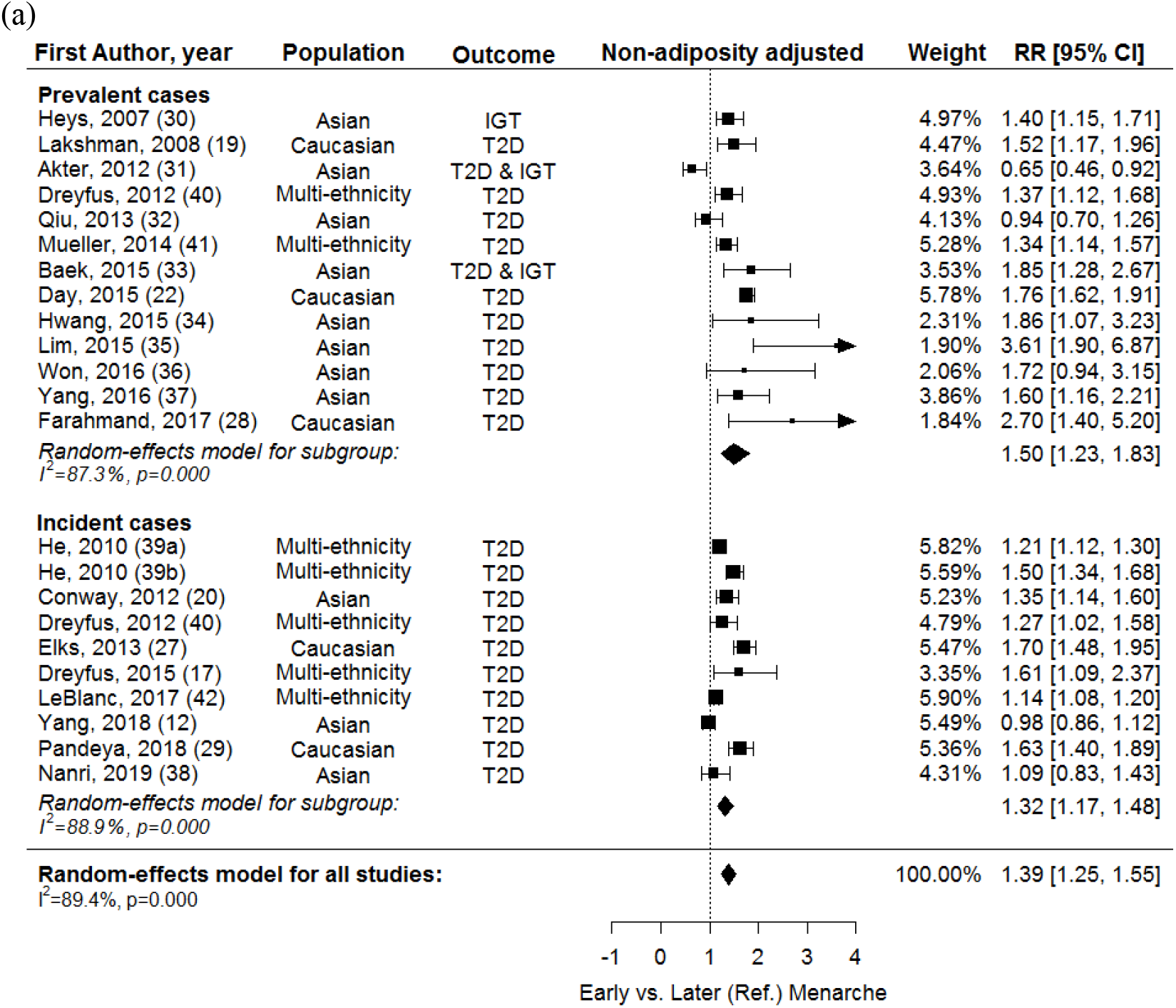
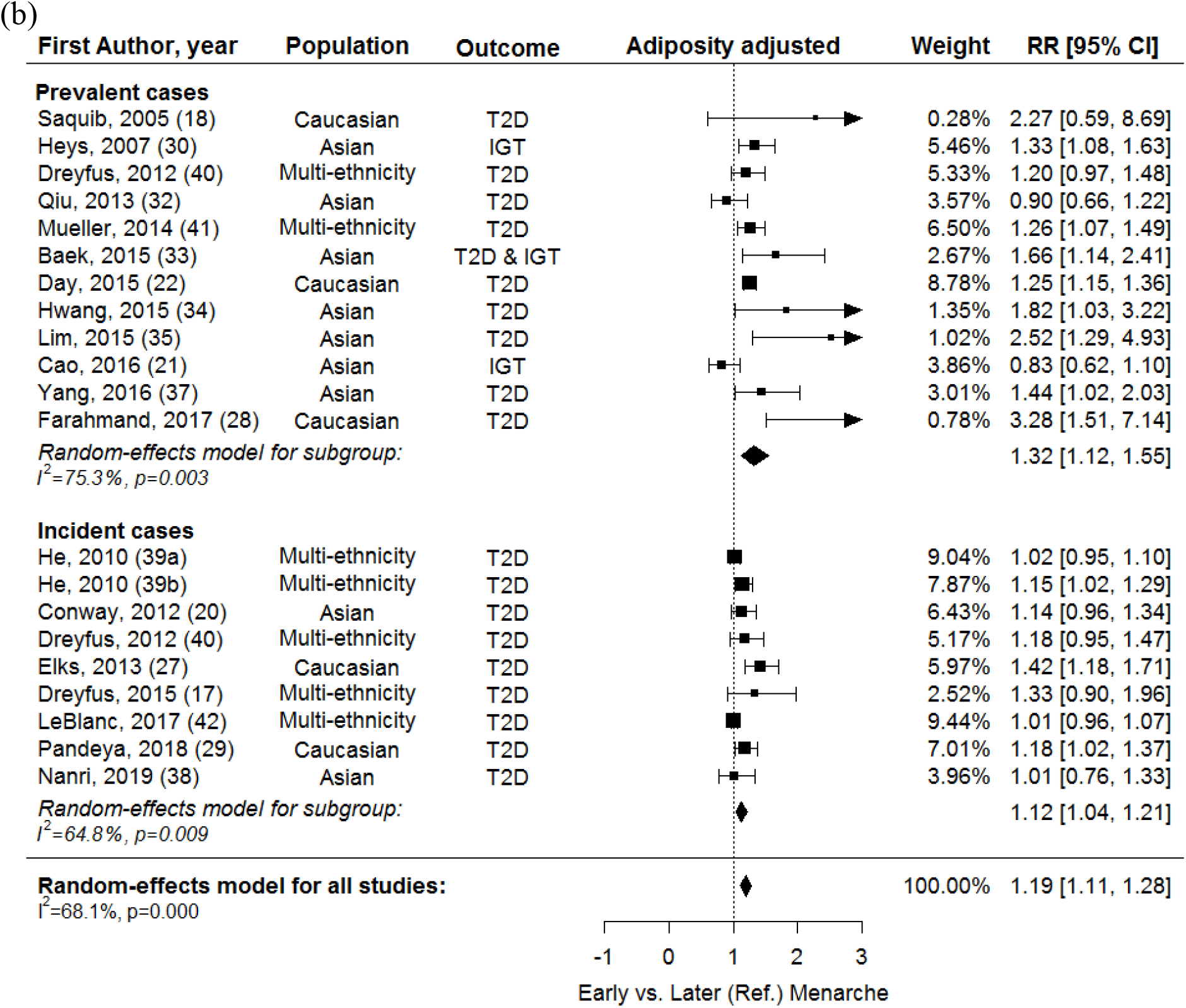
Forest plots of the association between early vs. later menarche and T2D/IGT, (a) without and (b) with adjustment for adiposity.

### Meta-regression results

Table 3 shows results of univariable meta-regression and pooled RR by subgroups of studies. Heterogeneity between studies was partially explained by ethnicity and study average AAM. The associations (continuous and categorical) between earlier menarche and higher T2D/IGT risk were stronger among studies of Caucasians than Asians, and stronger among populations with younger than older average AAM. In multivariable meta-regression analyses, only the contribution of study average AAM was evident, but not that of study ethnicity (data not shown). Year of enrolment, age at outcome assessment, number of variables adjusted, and the age cut-off used to define early menarche and the reference category did not explain the heterogeneity between study estimates (Supplemental Table 3).

**Table 3.** Univariable meta-regression and pooled RR for diabetes and glucose intolerance in subgroups.

| Factors | RR per year later age at menarche |  |  |  |  |  | Early versus later (Ref.) menarche |  |  |  |  |  |
| --- | --- | --- | --- | --- | --- | --- | --- | --- | --- | --- | --- | --- |
|  | Non-adiposity adjusted |  |  | Adiposity adjusted |  |  | Non-adiposity adjusted |  |  | Adiposity adjusted |  |  |
|  | N | RR (95% CI) | R<sup>2</sup> (%) | N | RR (95% CI) | R<sup>2</sup> (%) | N | RR (95% CI) | R<sup>2</sup> (%) | N | RR (95% CI) | R<sup>2</sup> (%) |
| Ethnicity |  |  | 53.7<sup>b</sup> |  |  | 99.63<sup>b</sup> |  |  | 52.3<sup>b</sup> |  |  | 16.2 |
| Asian | 3 | 0.95 (0.92, 0.97) |  | 2 | 0.98 (0.97, 0.99) |  | 11 | 1.37 (1.10, 1.71) |  | 9 | 1.23 (1.01, 1.49) |  |
| Caucasian | 5 | 0.88 (0.86, 0.90) |  | 6 | 0.95 (0.92, 0.98) |  | 5 | 1.72 (1.61, 1.83) |  | 5 | 1.27 (1.18, 1.36) |  |
| Multi-ethnic | 3 | 0.91 (0.87, 0.96) |  | 4 | 0.98 (0.96, 1.00) |  | 7 | 1.30 (1.18, 1.42) |  | 7 | 1.11 (1.02, 1.20) |  |
| Study average AAM, years<sup>a</sup> |  |  | 33.3 |  |  | 0 |  |  | 46.2 |  |  | 18.8 |
| <13.5 | 5 | 0.89 (0.86, 0.91) |  | 6 | 0.96 (0.93, 0.98) |  | 8 | 1.59 (1.45, 1.75) |  | 7 | 1.26 (1.19, 1.34) |  |
| ≥13.5 | 2 | 0.92 (0.85, 1.01) |  | 2 | 0.94 (0.86, 1.03) |  | 8 | 1.36 (1.17, 1.58) |  | 6 | 1.27 (1.03, 1.55) |  |
| *P value for linear trend* |  |  | <0.001<sup>b</sup> |  |  | 0.4 |  |  | 0.03<sup>b</sup> |  |  | 0.7 |

<sup>a</sup>Studies that did not report the information were excluded
<sup>b</sup>statistically significant in meta-regression: P<0.05
N, number of estimates, R<sup>2</sup> (%), % heterogeneity explained

### Assessment of publication bias and sensitivity analyses

Supplemental Figure 3 shows some asymmetry in funnel plots for studies on the categorical association between early menarche and T2D/IGT. Publication bias was statistically significant only for the studies on early vs. later menarche and T2D/IGT with adjustment for adiposity (Egger’s test, P<0.001).

Sensitivity analyses were performed to account for this publication bias. Supplemental Figure 4 shows the predicted missing studies using the trim-and-fill method. When the predicted missing studies were added to the meta-analyses, the continuous associations (adiposity unadjusted RR=0.91 per year, 95% CI=0.89-0.94; adiposity adjusted RR=0.97 per year, 95% CI=0.95-0.98) and categorical associations (adiposity unadjusted RR=1.35, 95% CI=1.21-1.49; adiposity adjusted RR=1.15, 95% CI=1.06-1.24) between earlier AAM and higher T2D/IGT risk remained similar.

Supplemental Figure 5 shows the results of leave-one-out analyses. When one of the study estimates was iteratively removed from the meta-analysis, the pooled estimates remained nearly unchanged for continuous and categorical associations between earlier AAM and higher T2D/IGT risk, with or without adjustment for adiposity.

### Contribution of early menarche to the burden T2D

The estimated population attributable risk for T2D/IGT due to early menarche among British women (<12 years; prevalence 20.15% in UK Biobank) unadjusted for adult adiposity was 12.6% (95% CI=11.0-14.3) and due to early menarche adjusted for adult adiposity was 5.1% (95% CI=3.6-6.7).

## DISCUSSION

The present meta-analysis of observational studies showed that earlier AAM is associated with higher T2D/IGT risk; this association is weaker but still evident after adjustment for adult adiposity. Study quality was in general high, and despite evidence of publication bias in one of the four models, similar findings were obtained in sensitivity analyses that considered predicted missing studies. Heterogeneity between studies was high and was partially explained by study differences in ethnicity and average AAM, with stronger associations in Caucasians and in study populations with lower average AAM. Assuming a causal relationship, a significant proportion of T2D/IGT among British women may be attributed to early menarche (before age 12 years). We found a paucity of studies on puberty timing and T2D/IGT in men.

Our meta-analysis findings are consistent with a previous review (11) but we included a larger number of studies (19 vs. 10) and women (890,583 vs. 315,428), we distinguished between findings unadjusted or adjusted for adiposity, and identified reasons for heterogeneity. While the previous meta-analysis (11) found the association of early menarche with higher T2D risk in Europe and the United States, we included more Asian studies and demonstrated that this association was also apparent in Asians, although weaker than in Caucasians, possibly due to their later average AAM. One study in China reported higher hazard ratios for incident diabetes associated with younger AAM in women born in the 1960s-1970s than in the 1950s and 1920s-1940s, consistent with the decreasing mean AAM from 16.2 years in 1920s-1940s to 14.7 years in 1960s-1970s (12). Hence, in light of worldwide secular trends towards declining average AAM (5,7-9), not only are more women moving into the high risk group (early menarche), but also the magnitude of elevated risk in this group appears to be increasing.

The mechanisms that underlie the association between earlier AAM and higher T2D/IGT risk are unclear. Early menarche is associated with rapid postnatal weight gain (44), and childhood (45,46) and adulthood obesity (47) which are known to be risk factors for T2D (48,49). However, our meta-analysis found that the association between earlier menarche and higher T2D/IGT risk remained, though attenuated, after accounting for potential confounding and mediating effects of adiposity, suggesting that there may be other adiposity-independent underlying mechanisms. It has been hypothesized that early menarche is the function of sex hormone exposure such as higher estradiol (50,51) and lower sex-hormone-binding globulin concentrations (52) in women, which may affect glycemic regulation and increase risk of diabetes (53-55). Nonetheless, hormone replacement therapy predominantly with estrogen was shown to reduce incidence of diabetes (56). Estrogen may have various effects on different parts of body including brain, adipose tissue, breast, endometrium and endothelium, probably mediated by different estrogen receptors (57).

We acknowledge several limitations of our study. We could not directly test or quantify the attenuation in the association when adjusting for adiposity, because the studies that contributed adjusted and unadjusted estimates were largely but not completely overlapping. All estimates were from observational studies and thus residual confounding may exist. AAM was mainly recalled during adulthood, which may affect its accuracy; however, moderate correlations between prospective and recalled AAM several decades later have been reported (58,59). Some publication bias was detected especially for the adiposity adjusted categorical association between early menarche and T2D/IGT, with potential bias towards reporting positive findings, although our sensitivity analyses were reassuring. The subgroup analyses by study average AAM were limited to studies that reported this value. Although we examined both continuous and categorical relationships between AAM and T2D/IGT risk, we were unable to examine if there was any threshold of AAM that indicates higher risk of T2D/IGT as indicated by one large study (27). Finally, we found only one study of puberty timing and T2D/IGT in men, likely because measures of puberty timing in men are not included in most studies. However, the one identified study was very large (n=197,714) and reported a statistically robust association between relatively younger (versus about average) voice breaking and T2D in men (adiposity unadjusted RR=1.44 (95% CI=1.30-1.59; adiposity adjusted RR=1.24 (95% CI=1.11-1.37)) (22).

In conclusion, observational studies show that earlier AAM is consistently associated with higher T2D/IGT risk, independent of adiposity. This association is stronger among Caucasians and populations with younger average AAM. Although the underlying mechanisms are not well understood, our summary findings quantify the potential benefits of avoiding early menarche to prevent T2D/IGT.

## Data Availability

Data are available from publications included in this manuscript

## Author Contributions

T.S.C. and K.K.O. contributed to study concept and design, acquisition of data, and drafting of the manuscript. T.S.C. contributed to statistical analysis of data. All authors contributed to interpretation of findings and to critical revision of the manuscript.

## Acknowledgement

We thank Stephen Sharp, MRC Epidemiology Unit, University of Cambridge, for statistical advice.

## References

1. Marshall WA, Tanner JM: Variations in the pattern of pubertal changes in boys. Archives of disease in childhood 1970;45:13–23

2. Marshall WA, Tanner JM: Variations in pattern of pubertal changes in girls. Archives of disease in childhood 1969;44:291–303

3. Ahmed ML, Ong KK, Dunger DB: Childhood obesity and the timing of puberty. Trends in Endocrinology & Metabolism 2009;20:237–242

4. Harries M, Walker JM, Williams DM, et al.: Changes in the male voice at puberty. Archives of disease in childhood 1997;77:445–447

5. Sorensen K, Mouritsen A, Aksglaede L, et al.: Recent secular trends in pubertal timing: implications for evaluation and diagnosis of precocious puberty. Hormone research in paediatrics 2012;77:137–145

6. Harrington J, Palmert MR: Clinical review: Distinguishing constitutional delay of growth and puberty from isolated hypogonadotropic hypogonadism: critical appraisal of available diagnostic tests. The Journal of clinical endocrinology and metabolism 2012;97:3056–3067

7. Hosokawa M, Imazeki S, Mizunuma H, et al.: Secular trends in age at menarche and time to establish regular menstrual cycling in Japanese women born between 1930 and 1985. BMC women’s health 2012;12:19–19

8. Cho GJ, Park HT, Shin JH, et al.: Age at menarche in a Korean population: secular trends and influencing factors. European journal of pediatrics 2010;169:89–94

9. van der Eng P, Sohn K: The biological standard of living in Indonesia during the 20th century: Evidence from the age at menarche. Economics and human biology 2018;

10. Kaiser AB, Zhang N, DER PLUIJM WV: Global Prevalence of Type 2 Diabetes over the Next Ten Years (2018-2028). Diabetes 2018;67:202–LB

11. Janghorbani M, Mansourian M, Hosseini E: Systematic review and meta-analysis of age at menarche and risk of type 2 diabetes. Acta diabetologica 2014;51:519–528

12. Yang L, Li L, Peters SAE, et al.: Age at Menarche and Incidence of Diabetes: A Prospective Study of 300,000 Women in China. American journal of epidemiology 2018;187:190–198

13. Au Yeung SL, Jiang C, Cheng KK, et al.: Age at menarche and cardiovascular risk factors using Mendelian randomization in the Guangzhou Biobank Cohort Study. Preventive medicine 2017;101:142–148

14. Evensen E, Wilsgaard T, Furberg AS, Skeie G: Tracking of overweight and obesity from early childhood to adolescence in a population-based cohort - the Tromso Study, Fit Futures. BMC pediatrics 2016;16:64

15. Freedman DS, Khan LK, Serdula MK, et al.: The relation of menarcheal age to obesity in childhood and adulthood: the Bogalusa heart study. BMC pediatrics 2003;3:3

16. Scottish Intercollegiate Guidelines Network [article online], Available from https://www.sign.ac.uk/search-filters.html.

17. Dreyfus J, Jacobs DR, Jr., Mueller N, et al.: Age at Menarche and Cardiometabolic Risk in Adulthood: The Coronary Artery Risk Development in Young Adults Study. The Journal of pediatrics 2015;167:344-352.e341

18. Saquib N, Kritz-Silverstein D, Barrett-Connor E: Age at menarche, abnormal glucose tolerance and type 2 diabetes mellitus: The Rancho Bernardo Study. Climacteric : the journal of the International Menopause Society 2005;8:76–82

19. Lakshman R, Forouhi N, Luben R, et al.: Association between age at menarche and risk of diabetes in adults: results from the EPIC-Norfolk cohort study. Diabetologia 2008;51:781–786

20. Conway BN, Shu XO, Zhang X, et al.: Age at menarche, the leg length to sitting height ratio, and risk of diabetes in middle-aged and elderly Chinese men and women. PloS one 2012;7:e30625

21. Cao X, Zhou J, Yuan H, Chen Z: Duration of reproductive lifespan and age at menarche in relation to metabolic syndrome in postmenopausal Chinese women. Journal of Obstetrics and Gynaecology Research 2016;42:1581–1587

22. Day FR, Elks CE, Murray A, et al.: Puberty timing associated with diabetes, cardiovascular disease and also diverse health outcomes in men and women: the UK Biobank study. Scientific reports 2015;5:11208

23. Stang A: Critical evaluation of the Newcastle-Ottawa scale for the assessment of the quality of nonrandomized studies in meta-analyses. European journal of epidemiology 2010;25:603–605

24. Pierce MB, Kuh D, Hardy R: The role of BMI across the life course in the relationship between age at menarche and diabetes, in a British Birth Cohort. Diabetic medicine : a journal of the British Diabetic Association 2012;29:600–603

25. Cooper GS, Ephross SA, Sandler DP: Menstrual patterns and risk of adult-onset diabetes mellitus. Journal of clinical epidemiology 2000;53:1170–1173

26. Stockl D, Doring A, Peters A, et al.: Age at menarche is associated with prediabetes and diabetes in women (aged 32-81 years) from the general population: the KORA F4 Study. Diabetologia 2012;55:681–688

27. Elks CE, Ong KK, Scott RA, et al.: Age at menarche and type 2 diabetes risk: the EPIC-InterAct study. Diabetes care 2013;36:3526–3534

28. Farahmand M, Tehrani FR, Dovom MR, Azizi F: Menarcheal Age and Risk of Type 2 Diabetes: A Community-Based Cohort Study. Journal of clinical research in pediatric endocrinology 2017;9:156–162

29. Pandeya N, Huxley RR, Chung HF, et al.: Female reproductive history and risk of type 2 diabetes: A prospective analysis of 126 721 women. Diabetes, obesity & metabolism 2018;20:2103–2112

30. Heys M, Schooling CM, Jiang C, et al.: Age of menarche and the metabolic syndrome in China. Epidemiology (Cambridge, Mass) 2007;18:740–746

31. Akter S, Jesmin S, Islam M, et al.: Association of age at menarche with metabolic syndrome and its components in rural Bangladeshi women. Nutrition & metabolism 2012;9:99

32. Qiu C, Chen H, Wen J, et al.: Associations between age at menarche and menopause with cardiovascular disease, diabetes, and osteoporosis in Chinese women. The Journal of clinical endocrinology and metabolism 2013;98:1612–1621

33. Baek TH, Lim NK, Kim MJ, et al.: Age at menarche and its association with dysglycemia in Korean middle-aged women. Menopause (New York, NY) 2015;22:542–548

34. Hwang E, Lee KW, Cho Y, et al.: Association between age at menarche and diabetes in Korean post-menopausal women: results from the Korea National Health and Nutrition Examination Survey (2007-2009). Endocrine journal 2015;62:897–905

35. Lim JS, Lee HS, Kim EY, et al.: Early menarche increases the risk of Type 2 diabetes in young and middle-aged Korean women. Diabetic medicine : a journal of the British Diabetic Association 2015;32:521–525

36. Won JC, Hong JW, Noh JH, Kim DJ: Association Between Age at Menarche and Risk Factors for Cardiovascular Diseases in Korean Women: The 2010 to 2013 Korea National Health and Nutrition Examination Survey. Medicine 2016;95:e3580

37. Yang A, Liu S, Cheng N, et al.: Reproductive factors and risk of type 2 diabetes in an occupational cohort of Chinese women. Journal of diabetes and its complications 2016;30:1217–1222

38. Nanri A, Mizoue T, Noda M, et al.: Menstrual and reproductive factors and type 2 diabetes risk: The Japan Public Health Center-based Prospective Study. Journal of diabetes investigation 2019;10:147–153

39. He C, Zhang C, Hunter DJ, et al.: Age at menarche and risk of type 2 diabetes: results from 2 large prospective cohort studies. American journal of epidemiology 2010;171:334–344

40. Dreyfus JG, Lutsey PL, Huxley R, et al.: Age at menarche and risk of type 2 diabetes among African-American and white women in the Atherosclerosis Risk in Communities (ARIC) study. Diabetologia 2012;55:2371–2380

41. Mueller NT, Duncan BB, Barreto SM, et al.: Earlier age at menarche is associated with higher diabetes risk and cardiometabolic disease risk factors in Brazilian adults: Brazilian Longitudinal Study of Adult Health (ELSA-Brasil). Cardiovascular diabetology 2014;13:22

42. LeBlanc ES, Kapphahn K, Hedlin H, et al.: Reproductive history and risk of type 2 diabetes mellitus in postmenopausal women: findings from the Women’s Health Initiative. Menopause (New York, NY) 2017;24:64–72

43. Petersohn I, Zarate-Ortiz AG, Cepeda-Lopez AC, Melse-Boonstra A: Time Trends in Age at Menarche and Related Non-Communicable Disease Risk during the 20th Century in Mexico. Nutrients 2019;11

44. Terry MB, Ferris JS, Tehranifar P, et al.: Birth Weight, Postnatal Growth, and Age at Menarche. American journal of epidemiology 2009;170:72–79

45. Li W, Liu Q, Deng X, et al.: Association between Obesity and Puberty Timing: A Systematic Review and Meta-Analysis. International journal of environmental research and public health 2017;14:1266

46. Mumby HS, Elks CE, Li S, et al.: Mendelian Randomisation Study of Childhood BMI and Early Menarche. Journal of obesity 2011;2011:180729

47. Trikudanathan S, Pedley A, Massaro JM, et al.: Association of female reproductive factors with body composition: the Framingham Heart Study. The Journal of clinical endocrinology and metabolism 2013;98:236–244

48. Al-Goblan AS, Al-Alfi MA, Khan MZ: Mechanism linking diabetes mellitus and obesity. Diabetes, metabolic syndrome and obesity : targets and therapy 2014;7:587–591

49. Eriksson JG, Forsen TJ, Osmond C, Barker DJP: Pathways of Infant and Childhood Growth That Lead to Type 2 Diabetes. Diabetes care 2003;26:3006–3010

50. Vihko R, Apter D: Endocrine characteristics of adolescent menstrual cycles: impact of early menarche. Journal of steroid biochemistry 1984;20:231–236

51. Apter D, Vihko R: Early menarche, a risk factor for breast cancer, indicates early onset of ovulatory cycles. The Journal of clinical endocrinology and metabolism 1983;57:82–86

52. Thankamony A, Ong KK, Ahmed ML, et al.: Higher levels of IGF-I and adrenal androgens at age 8 years are associated with earlier age at menarche in girls. The Journal of clinical endocrinology and metabolism 2012;97:E786–790

53. Ding EL, Song Y, Malik VS, Liu S: Sex differences of endogenous sex hormones and risk of type 2 diabetes: a systematic review and meta-analysis. Jama 2006;295:1288–1299

54. O’Reilly MW, Glisic M, Kumarendran B, et al.: Serum testosterone, sex hormone-binding globulin and sex-specific risk of incident type 2 diabetes in a retrospective primary care cohort. Clinical endocrinology 2019;90:145–154

55. Perry JR, Weedon MN, Langenberg C, et al.: Genetic evidence that raised sex hormone binding globulin (SHBG) levels reduce the risk of type 2 diabetes. Human molecular genetics 2010;19:535–544

56. Salpeter SR, Walsh JM, Ormiston TM, et al.: Meta-analysis: effect of hormone-replacement therapy on components of the metabolic syndrome in postmenopausal women. Diabetes, obesity & metabolism 2006;8:538–554

57. Clegg D, Hevener AL, Moreau KL, et al.: Sex Hormones and Cardiometabolic Health: Role of Estrogen and Estrogen Receptors. Endocrinology 2017;158:1095–1105

58. Must A, Phillips SM, Naumova EN, et al.: Recall of early menstrual history and menarcheal body size: after 30 years, how well do women remember? American journal of epidemiology 2002;155:672–679

59. Cooper R, Blell M, Hardy R, et al.: Validity of age at menarche self-reported in adulthood. Journal of epidemiology and community health 2006;60:993–997

